# Is 7p14.1 an orofacial cleft risk locus? Genome-wide study of copy number variation in multiple populations provides both a replication of previous studies and an alternative explanation

**DOI:** 10.64898/2026.01.09.26343782

**Authors:** Nandita Mukhopadhyay, Eleanor E. Feingold, Harrison Brand, Myoung Keun Lee, Edibe Nehir Kurtas, Alba Sanchis-Juan, Lina Moreno-Uribe, George Wehby, Luz Consuelo Valencia-Ramirez, Claudia P. Restrepo Muñeton, Carmencita Padilla, Frederic Deleyiannis, Fernando A. Poletta, Ieda M. Orioli, Jacqueline T. Hecht, Carmen J. Buxó, Azeez Butali, Wasiu L. Adeyemo, Mekonen Eshete Abebe, Alexandre R. Vieira, John R. Shaffer, Jeffrey C. Murray, Seth M. Weinberg, Ingo Ruczinski, Elizabeth J. Leslie-Clarkson, Mary L. Marazita

**Affiliations:** Center for Craniofacial and Dental Genetics, Department of Oral and Craniofacial Sciences, School of Dental Medicine, University of Pittsburgh, Pittsburgh, PA, 15219 USA; College of Science, Oregon State University; Center for Genomic Medicine, Mass General Research Institute, Boston, MA 02114; Department of Neurology, Harvard Medical School, Boston, MA, USA; Department of Surgery, Harvard Medical School, Boston, MA, USA; Department of Orthodontics, & The Iowa Institute for Oral Health Research, College of Dentistry, University of Iowa, Iowa City, IA, USA; Department of Health Management and Policy, College of Public Health, University of Iowa, Iowa City, IA, USA; Fundación Clínica Noel; Calle 14 # 43B – 146, Medellín, Antioquia, Colombia; Department of Pediatrics, College of Medicine, Institute of Human Genetics, National Institutes of Health, University of the Philippines, Manila, the Philippines; UCHealth Medical Group, Colorado Springs, CO. USA; CEMIC-CONICET: Center for Medical Education and Clinical Research, Buenos Aires, Argentina; Department of Genetics, Institute of Biology, Federal University of Rio de Janeiro, Rio de Janeiro, Brazil; Instituto Nacional de Genética Médica Populacional INAGEMP, Porto Alegre, Brazil; Department of Pediatrics, University of Texas Health Science Center at Houston, Houston, TX, USA; Dental and Craniofacial Genomics Core, School of Dental Medicine, University of Puerto Rico, San Juan, Puerto Rico; Department of Oral Pathology, Radiology and Medicine and Iowa Institute for Oral Health Research, College of Dentistry, University of Iowa, Iowa City, IA, USA; Department of Oral and Maxillofacial Surgery, College of Medicine, University of Lagos, Lagos, Nigeria; Department of Plastic and Reconstructive Surgery, Addis Ababa University, Ethiopia; East Carolina University, School of Dental Medicine, Greenville, NC 27834-4354; Department of Human Genetics, School of Public Health, University of Pittsburgh, Pittsburgh, PA, USA; Department of Pediatrics, Carver College of Medicine, University of Iowa, Iowa City, IA, USA; Department of Biostatistics, Johns Hopkins School of Public Health. 615 North Wolfe Street, Baltimore, MD 21205-2179; Department of Human Genetics, Emory University School of Medicine, Atlanta, GA, USA; Clinical and Translational Science, School of Medicine, University of Pittsburgh, Pittsburgh, PA, USA

**Keywords:** Copy number variation, orofacial cleft, genomewide association, family study

## Abstract

**Objective:** Our understanding of the genetic causes of non-syndromic orofacial clefts (OFCs) is based largely upon genetic studies of common and rare nucleotide variants. Less is known about the role of copy number variations (CNVs) and the studies published to date have been limited to either small samples or targeted genomic regions. The objective of our study is to investigate the contribution of CNVs spread across the entire genome to OFC risk in a large multi-ancestry cohort.

**Methods:** We utilized PennCNV on microarray genotyping data to detect CNVs in 10,240 participants (2,484 with clefts, 7,756 unaffected). 70,695 quality-filtered autosomal CNVs (49,660 deletions, 21,035 duplications) were used to assign normal/abnormal copy number statuses at 67,199 positions from the GRCh37 genome assembly. Genome-wide association was run between cleft status and copy number status.

**Results:** We observed a highly significant association between OFCs and deletions on chromosome 7p14.1 (p=1.32e-35) driven by Central and South American ancestry (p=1.04e-25) participants, with less significant contributions from European (p=3.37e-08) and Asian (p=0.01) ancestry participants. We also observed four other loci with *p*-values below 10e-04.

**Conclusion:** The 7p14.1 association observed in our study is a replication of two prior studies in independent cohorts of European ancestry. However, this locus lies in a T-cell receptor region that is subject to somatic rearrangements that decrease in frequency with age and may affect genetic association results. Our data show age effects as well as differences between blood and saliva samples. Thus, our results can be interpreted either as supporting a previously established association with orofacial clefts, or as questioning those previous results in favor of a hypothesis about the behavior of somatic rearrangements in T-cell receptor regions.

## INTRODUCTION

Orofacial clefts (OFCs) are the most prevalent craniofacial anomalies in humans, affecting approximately 1 in 700 newborns worldwide, and are thus one of the most common structural birth defects. A child with an OFC initially faces feeding difficulties and undergoes multiple surgeries and years of orthodontic and speech therapy, which poses a significant social, emotional, and financial burden for individuals affected with OFCs, their families, and society (Berk & Marazita, 2002; Naros et al., 2018; Semb et al., 2005; Wehby & Cassell, 2010). Further, individuals born with an OFC have an increased incidence of mental health problems, higher risk for other disorders including breast, brain, and colon cancers, as well as higher mortality at all stages of life (particularly in developing countries where access to medical care may be limited) (C. Bille et al., 2005; Camilla Bille et al., 2005; Christensen et al., 2004; Dietz et al., 2012a, 2012b; Menezes et al., 2009; Wehby & Cassell, 2010; Wehby et al., 2006; Zhu et al., 2002). The impact of OFCs is further complicated by differences in birth prevalence among human populations, ranging from ∼1/500 live births in Native American and Asian populations to ∼1/2500 in African populations (Christensen, 1999; Cooper et al., 2006; Mossey, 2007; Mossey et al., 2009). Although treatments such as surgical remediation, ongoing orthodontia, speech and other therapies are available, these are costly and differ in terms of accessibility. Thus, identifying OFC etiologic factors is vital for assessing risk, the development of prevention methods, and determining the extent of therapeutic and social support needed by affected individuals and their families.

Phenotypically, OFCs vary by manifestation and severity, being broadly classified as three subtypes: cleft lip alone (CL), cleft palate alone (CP), and cleft lip plus cleft palate (CLP). Based on recurrence and etiology, CL and CLP are frequency combined as cleft lip with or without cleft palate (CL/P). The majority occur in isolation, i.e. without any other detectable cognitive or structural abnormality (Dixon et al., 2011) - about 70% CL/P and 50% of CP alone. Our understanding of the genetic causes of non-syndromic OFCs has made enormous progress over the last few decades but is still incomplete. The current picture of the genetic architecture of OFCs is based largely on the dozens of genome-wide studies published over the last 15 years of the contribution of SNPs from genotyping arrays and sequencing (see review by Alade et al., 2022). However, much less is known about the role of other classes of genetic variants, such as copy number variants (CNVs).

Several prior studies have investigated the contribution of CNVs to OFC risk. Most of these were focused only on known CNV regions and were limited by the sample size and/or target regions analyzed (Conte et al., 2016; Lansdon et al., 2023; Simioni et al., 2015). The Simioni et al study looked at CNVs 300 KB or longer within 8 OFC risk genes in 23 unrelated individuals with isolated OFCs and OFCs in conjunction with other anomalies using array CGH, and concluded that CNVs could not explain the OFC phenotypes. Conte et al. conducted a gene enrichment analysis of 249 deletions and 226 duplications present in 312 DECIPHER and ECARUCA participants with OFCs. CNVs present in 2 or more participants were observed in 2 OFC risk genes and 12 others involved in craniofacial enrichment. Lansdon et al. looked at CNVs overlapping Mendelian OFC genes in 869 Filippino syndromic and non-syndromic OFC patients from Operation using array CGH to identify CNVs. They found no overall differences in CNV burden between non-syndromic and syndromic OFC patients, nor by cleft type within the non-syndromic cleft group. In contrast, studies by Younkin et al. (Younkin et al., 2014, 2015) were conducted on a substantially larger sample (467 CL/P case and 391 control trios) of white European ancestry utilizing a GWAS SNP array to analyze *de novo* and inherited deletions separately and found a region within 7p14.1 to be enriched for deletions within case trios. This locus was subsequently replicated by Klamt et al (Klamt et al., 2016) using a case-control study of 399 cases and 1,218 controls also of European ancestry.

The current study investigated whether abnormal copy number, i.e. the presence of a deletion or duplication, is associated with non-syndromic CL/P, using CNVs identified from genotype intensities in a GWAS SNP panel from the multiethnic Pittsburgh Orofacial Cleft Study (POFC). A genome-wide case-control association framework was then used to analyze the presence/absence of CNVs at predetermined genomic coordinates located across the genome. In contrast to prior CNV studies, our approach is more general in nature – both *de novo* and inherited CNVs are treated similarly, and for the most part, we combined analysis of deletions and duplications under the assumption that both may be similarly disruptive to gene functioning. Further, our statistical method, which tests association between phenotype and CNV status at pre-determined positions rather than for specific CNVs, also reduces the need to accurately resolve breakpoints or to determine whether two separately called CNVs are in fact distinct from each other.

## SUBJECTS AND METHODS

### Sample

The sample analyzed in this study consists of participants from the multi-ancestry Pittsburgh Orofacial Cleft study (POFC), including a variety of simplex and multiplex pedigrees of varying sizes and geographic origin. Study participants were recruited with the approval of the Pittsburgh coordinating center, and site-specific ethics committees. Genotyping was conducted at the Center for Inherited Disease Research (CIDR) at Johns Hopkins University, for approximately 580,000 genome-wide variants. Assessment of genotyping quality was performed by the CIDR coordinating center at the University of Washington. Detailed descriptions of sample collection and genotyping procedures are available in previous publications (Leslie et al., 2017; Leslie et al., 2016). Subject phenotype and genotype data are available from dbGaP (dbGaP Study Accession numbers: phs000774.v2.p1; phs000440.v1.p1).

The sample utilized in our current study includes 2,484 individuals affected with CL or CLP and 5,264 unaffected relatives from 2,258 families that contain members affected with CL and/or CLP. An additional 2,492 unaffected individuals from 1,422 families with no observed OFCs, nor a reported history of OFCs in relatives within the 3^rd^ degree, were also included in the study sample as controls. Participants who were affected with CP and their relatives were excluded from this study.

The study sample was partitioned into four genetically-defined ancestry groups, using principal components analysis defined in a previous study (Leslie et al., 2016), without taking into account self-reported race or ancestry. These four ancestry groups are identified as African, Asian, European, and Central and South American origin, and the partitioning largely follows the geographic locations of recruitment sites, except for participants from recruitment sites within the US, who are represented in multiple groups. Table 1 below lists sites represented within each group.

**Table 1.** Subgroups based on genetic ancestry and recruitment sites within the combined study sample.

| **Group** | **Site** | **Count** |
| --- | --- | --- |
| Central and South American | Colombia | 2,361 |
|  | Argentina | 427 |
|  | Guatemala | 814 |
|  | Puerto Rico | 412 |
|  | United States | 350 |
|  | **Subtotal Central and South American Sites** | **4,364** |
| European | United States | 1,519 |
|  | Hungary | 942 |
|  | Turkey | 599 |
|  | Denmark | 300 |
|  | Spain | 119 |
|  | **Subtotal European sites** | **3,479** |
| Asian | Philippines | 855 |
|  | China | 794 |
|  | India | 332 |
|  | **Subtotal Asian sites** | **1,981** |
| African | Nigeria | 184 |
|  | Ethiopia | 162 |
|  | United States | 70 |
|  | **Subtotal African sites** | **416** |
| **All groups** | **All Sites** | **10,240** |

### Identification and filtering of CNVs

The CNVs were called using the program PennCNV (version 1.0.0) (Wang et al., 2007), within each of the four ancestry groups. Subsequently, association analysis was run within each group separately in addition to the pooled association analysis of all POFC participants.

PennCNV uses a hidden Markov model (HMM) to detect copy number variation using data from a high-density SNP genotyping array. For our study, we used a previously published exome-based HMM model based on a large Swedish population for the Illumina HumanCoreExome_v12-A beadchip array (Szatkiewicz et al., 2013). Population-based B allele frequencies were separately compiled using the B-allele frequency values of control subjects from families without observed or a history of OFCs corresponding to each ancestry, referred to in the rest of this report as true controls. For most of the analyses, the unaffected family members and true controls are analyzed as a single group – however, in a few cases, we report results separately for these two groups.

CNV calls within centromeric and telomeric regions were filtered out. However, we also observed excessive numbers of CNV calls spanning the ends of each chromosome within the remaining CNV calls (Supplement Figure 1). Therefore, regions measuring approximately 5% of the chromosome length located at the ends each chromosome were also excluded from our analyses. The GC-adjusted CNVs within the retained regions of the genome were quality filtered based on the log likelihood ratios observed within each CNV region; individual CNVs with a large standard deviation (> 0.3) in log R ratios were removed. Adjacent CNVs were merged into longer segments if the gap between the two segments was smaller than 1/5 the total span of the two CNVs. Merged CNVs that spanned fewer than 10 call sites as well as those longer than 5 MB were also ignored, and copy numbers set to normal within these CNV calls.

Prior to running a CNV GWAS, we tested whether there were differences in the mean number of variants per individual due to their sex and ancestry by carrying out an analysis-of-variance analysis.

### Genome-wide Association

A CNV based genome-wide association against CL/P status was run in three ways: 1) abnormal copy number including deletions and duplications (DEL+DUP) vs. normal copy number, 2) deletions only (DEL) vs. normal copy number, and 3) duplications only (DUP) vs. normal copy number. For each analysis, the model fit the outcome (affected with a CL or a CLP, or unaffected) as a function of copy number along with sex as the other predictor variable.

Because the dataset as a whole contains relatively small numbers of duplications, this third analysis was only performed as a follow-up analysis for regions of interest based on the first two analyses.

Each of the analyses described above was carried out position by position across the genome using ∼580K SNP positions from the GRCh37 genome assembly on the HumanCoreExome_v12-A beadchip array. At a given position, each person was scored as “normal copy number,” “duplication,” or “deletion”. The statistical analyses compared the frequency of abnormal copy number between CL/P affected and unaffected participants at each location. This approach is in contrast to typical CNV association studies that attempt to identify shared CNVs among samples. By doing analyses location by location, our approach generates association statistics that form broad peaks across CNV regions, with the smallest p-values typically occurring at the location(s) where the greatest number of samples have abnormal copy number. This technique has been previously used to study association of CNVs to psychosis in Alzheimer’s disease (Zheng et al., 2015). Figure 1 outlines the procedure we followed for running genome-wide analysis of copy-number in the POFC study participants.

**Figure 1.**
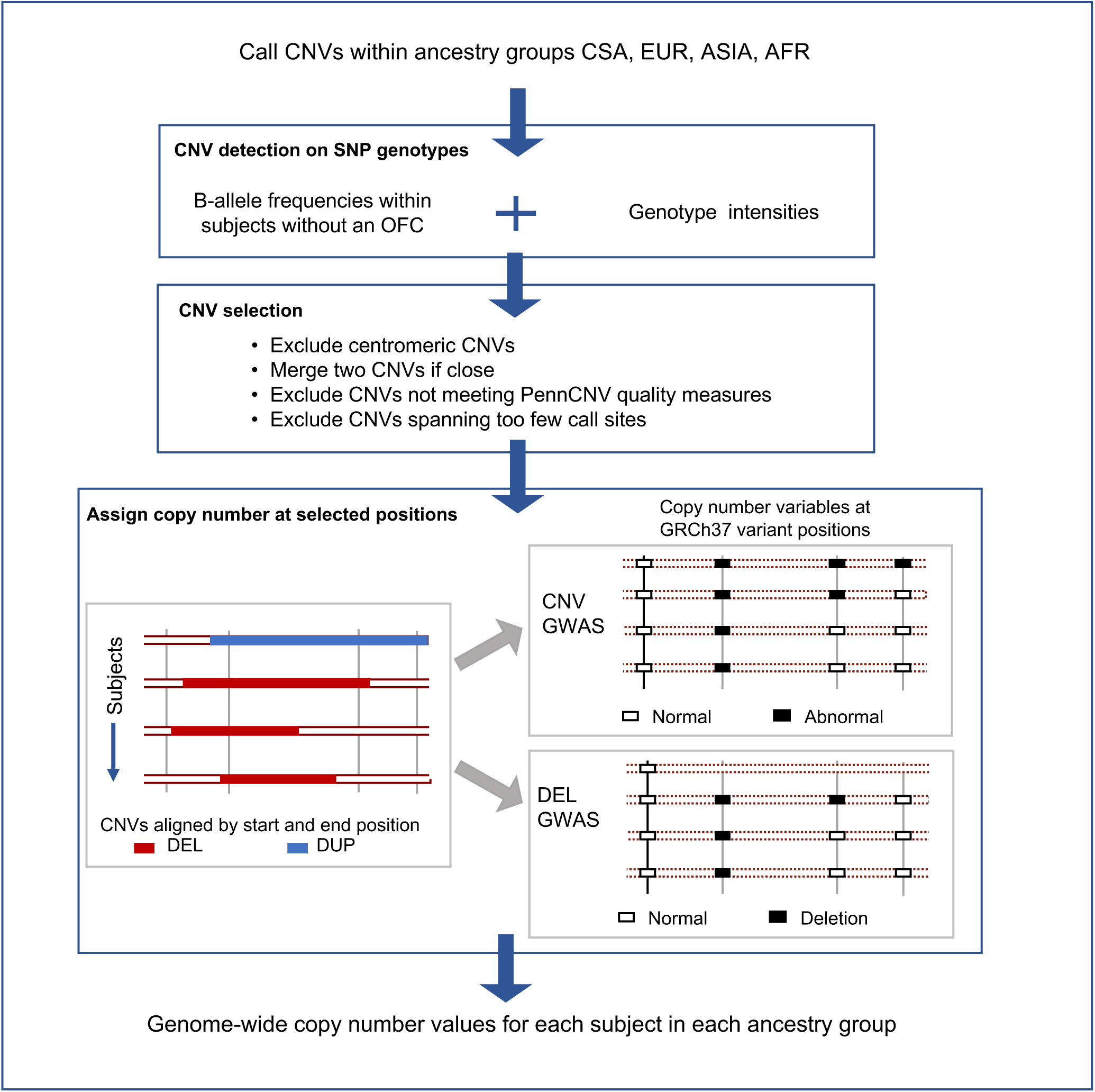
CNV scoring and derivation of copy number variables for GWAS. Schematic representation of CNV identification within four ancestry-based subgroups in the POFC study, followed by quality filtering and merging of smaller adjacent CNVs into larger contiguous CNVs, and assignment of normal/abnormal copy number at pre-determined positions from GRcH 37 for genome-wide association analysis.

Associations were run on the combined sample and in three of the ancestry subgroups described above, Central and South Americans, Europeans and Asians, the African sample size being too small to analyze alone. Individuals with a cleft lip (CL) or a cleft lip with cleft palate (CLP) were scored as affected; family members without CL or CLP and controls from families without observed OFCs or a history of OFCs were scored as unaffected. We used EMMAX, a variance component-based case-control association framework using a genetic relationship matrix (GRM) to account for relatedness and population admixture. Our binary copy number variable was treated as if it were a dominant genotype. That is, normal copy number was coded as zero and abnormal copy number was coded as 1. The GRM in this study was computed previously using imputed SNP genotype data, under EMMAX’s kinship estimation (Kang et al., 2010). The EMMAX association method provides only approximate estimates of effect size for association of a binary phenotype, therefore we only report the direction of the main effect in the results below. For all analyses, we filtered out genomic positions where fewer than 10 individuals had abnormal copy number.

### Discovery and investigation of associated regions

In order to set a statistical significance standard, we noted that the CNV statuses at adjacent positions are highly correlated due to the nature of CNVs which necessarily span multiple positions. Therefore, the observed association p-values are expected to occur in blocks, similar to those formed by linkage disequilibrium in a traditional SNP GWAS, but here, the blocks are substantially longer. Therefore, we set a pvalue threshold of 1e-04 for identifying noteworthy associations based on the total number of positions tested (94,468 sites located within one or more CNVs) divided by 10, the minimum number of sites spanned by CNVs used in GWAS.

We identified peaks in the combined sample and ancestry-based GWASs using our threshold. Each peak’s interval (i.e. leftmost and rightmost positions with the most significant p-values) was extended by 50 kb to either side, and we compared the association p-values in each such interval across all GWASs to identify loci that showed shared association across ancestry groups and CNV types.

CNVs located within associated regions were further examined to determine whether CNV sizes differed by affection status, ancestry, sex and whether they were inherited or *de novo*. Parent-offspring trios were selected from the cohort and the offspring’s CNV was judged to be a *de novo* only if neither parent had a CNV that overlapped the offspring’s CNV. For apparent *de novo* CNVs, we compared Log R ratios and B allele frequencies of offspring and their parents.

## RESULTS

### Overall CNV distribution

A total of 70,695 unique quality-filtered CNV regions were retained for genome-wide association. These consist of 49,660 deletions and 21,035 duplications. The CNVs range in length between 17 bp and 4,963,768 bp with median length 41,278 bp and 90% of CNVs ranging between 2,435-348,505 bp. Below we describe the frequency of CNVs based on different properties of the genome and the study sample (Table 2). Table 2 shows the breakdown of number of CNVs, DELs and DUPs by ancestry and affection status of the individuals, and the results of our comparison are discussed below.

**Table 2.** Individuals with abnormal copy number by CL/P status and ancestry after excluding CNVs at the ends of chromosomes.

| Ancestry | N<sub>SAMPLE</sub> | CNVs in study |  | DELs in study |  | DUPs in study |  | CNVs in CL/P |  |  | CNVs in Unaffected |  |  |
| --- | --- | --- | --- | --- | --- | --- | --- | --- | --- | --- | --- | --- | --- |
|  |  | Total | Mean per person (SD) | Total | Mean per person (SD) | Total | Mean per person (SD) | N<sub>SAMPLE</sub> | N<sub>CNV</sub> | Mean SD | N<sub>SAMPLE</sub> | N<sub>CNV</sub> | Mean SD |
| Central and South American sample | 4,364 | 126,617 | 29.04 (65.33) | 92,433 | 21.18 (62.21) | 34,284 | 7.86 (20.92) | 1,169 | 32,869 | 28.12 59.09 | 3,195 | 93,848 | 29.37 65.47 |
| European sample | 3,479 | 142,332 | 40.91 70.78 | 93,921 | 27.00 (64.65) | 48,411 | 13.92 (32.68) | 651 | 27,956 | 42.94 75.82 | 2,828 | 114,376 | 40.44 69.57 |
| Asian sample | 1,981 | 95,744 | 48.33 76.00 | 21,264 | 10.73 (44.72) | 74,480 | 37.60 (61.42) | 518 | 24,740 | 48.53 77.78 | 1,463 | 71,004 | 48.53 75.39 |
| African sample | 416 | 11,804 | 28.38 47.64 | 6,004 | 14.62 (41.74) | 5,720 | 13.75 (24.54) | 146 | 3,963 | 27.14 59.09 | 270 | 7,841 | 29.04 40.22 |
| Combined sample | 10,240 | 376,597 | 36.78 69.20 | 213,702 | 20.87 (59.67) | 162,895 | 15.91 (37.73) | 2,484 | 89,528 | 36.11 71.06 | 7,756 | 287,069 | 37.01 68.65 |

### Distribution by chromosome

We found different frequencies of CNVs per chromosome as measured by the total number of CNVs observed divided by the length of the chromosome in megabases. Chromosome 19 had double the frequency of CNVs (520 CNVs per MB) of chromosome 22, which had the second highest frequency (243 CNVs per MB), while chromosomes 13 and 18 had a substantially lower frequency of CNVs (75 and 80 per MB respectively). Supplement figure 2 shows the frequency of CNVs per MB by chromosome.

### Sex and ancestry

We noted statistically significant differences in number of CNVs, DELs and DUPs per person by ancestry (p-values for all three comparisons < 2.0e-16), with the highest frequency of CNVs observed in the Asian sample, followed by Europeans, Central and South Americans and Africans in order. Table 2 shows the mean number of CNVs, DELs and DUPs per individual for each ancestry. Although the average number of CNVs did not differ significantly between males and females (38 and 36 respectively, p=0.20), the average number of deletions per person was higher in females compared to males (22 and 19 respectively, p=0.006), whereas the opposite was true for duplications (15 and 17 respectively, p=0.04).

### Affection status

The mean number of CNVs (i.e. deletions and duplications combined) was not significantly different between CL/P-affected and unaffected participants (36 vs. 37, p-value = 0.20). Neither was the number of deletions and duplications, when considered separately (p-value = 0.68 and 0.64, respectively). We also looked at differences between CL/P cases, unaffected relatives of CL/P cases and true controls separately, in contrast to the earlier comparison that combine both types of unaffected participants into a single group. The mean number of CNVs in unaffected relatives of CL/P affected participants differed significantly from the true controls (38 vs. 34, p-value 0.041), whereas CL/P cases do not differ significantly from true controls (36 vs. 34, p-value 0.63). Based on these observations, we cannot conclude that, in this study, the risk of OFC in an individual is attributable to a polygenic genome-wide increased burden of CNVs.

### Genome-wide association of copy number statuses and CL/P

To test the hypothesis that genomic regions with abnormal copy number increase risk for CL/P, we ran a GWAS of abnormal copy number (deletions or duplications, DEL+DUP) and a second GWAS of deletions only (DEL). Five loci (1q42.13, 7p14.1, 10p12.1, 12q21.1, 22q11.21) exceeded a significance level of 10e-04 in the combined sample (Figure 2) and an additional three loci were significant in the European sample (1p35.2 and 19p13.11 in the DEL+DUP GWAS, and 21q22.3 in the DEL GWAS). Supplement Table 1 shows the p-value, position and number of CNVs and deletions for each peak. All effect directions were positive indicating that the presence of abnormal copy number or deletions was associated with increased risk of CL/P. The type-1 error rate of the genome-wide p-values did not appear to be inflated overall (see Supplement Figure 3 for QQ plots with and without the 7p14.1 locus). A detailed exploration of CNVs calls within the 7p14.1 locus is presented below.

**Figure 2.**
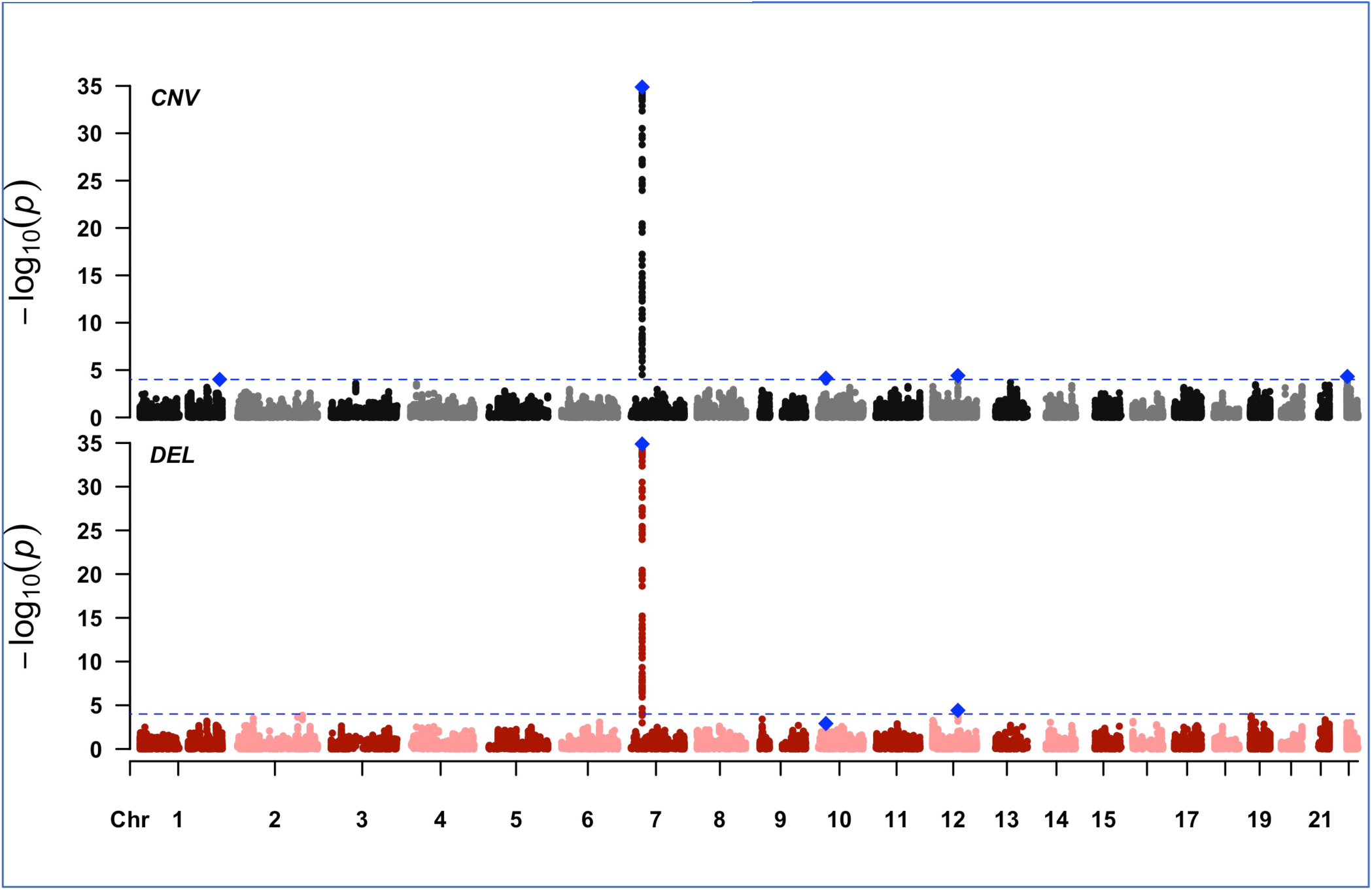
GWASs of CL/P in TOTAL population. Top panel shows GWAS of CNVs in the TOTAL population, using absence/presence of abnormal copy number, and bottom panel shows GWAS of deletions in the TOTAL population, using presence/absence of copy number < 2 at pre-determined GRcH 37 positions. Peaks with association p-values :-s 10e-04 are highlighted in blue, and the dotted line represents the significance threshold of 10e-04.

### Association at the 7p14.1 locus

The strong association observed at the 7p14.1 locus was attributable entirely to deletions. The most significant p-value was observed at 38,329,818 bp (association (p=1.32e-35, positive main effect) and was spanned by 196 total deletions (a frequency of approximately 2%). The frequency of deletions in affected participants was 8%, consistent with strong association seen at this locus. No duplications were seen within 100 kb of the peak in any of the four ancestry subgroups (Table 3).

**Table 3.**
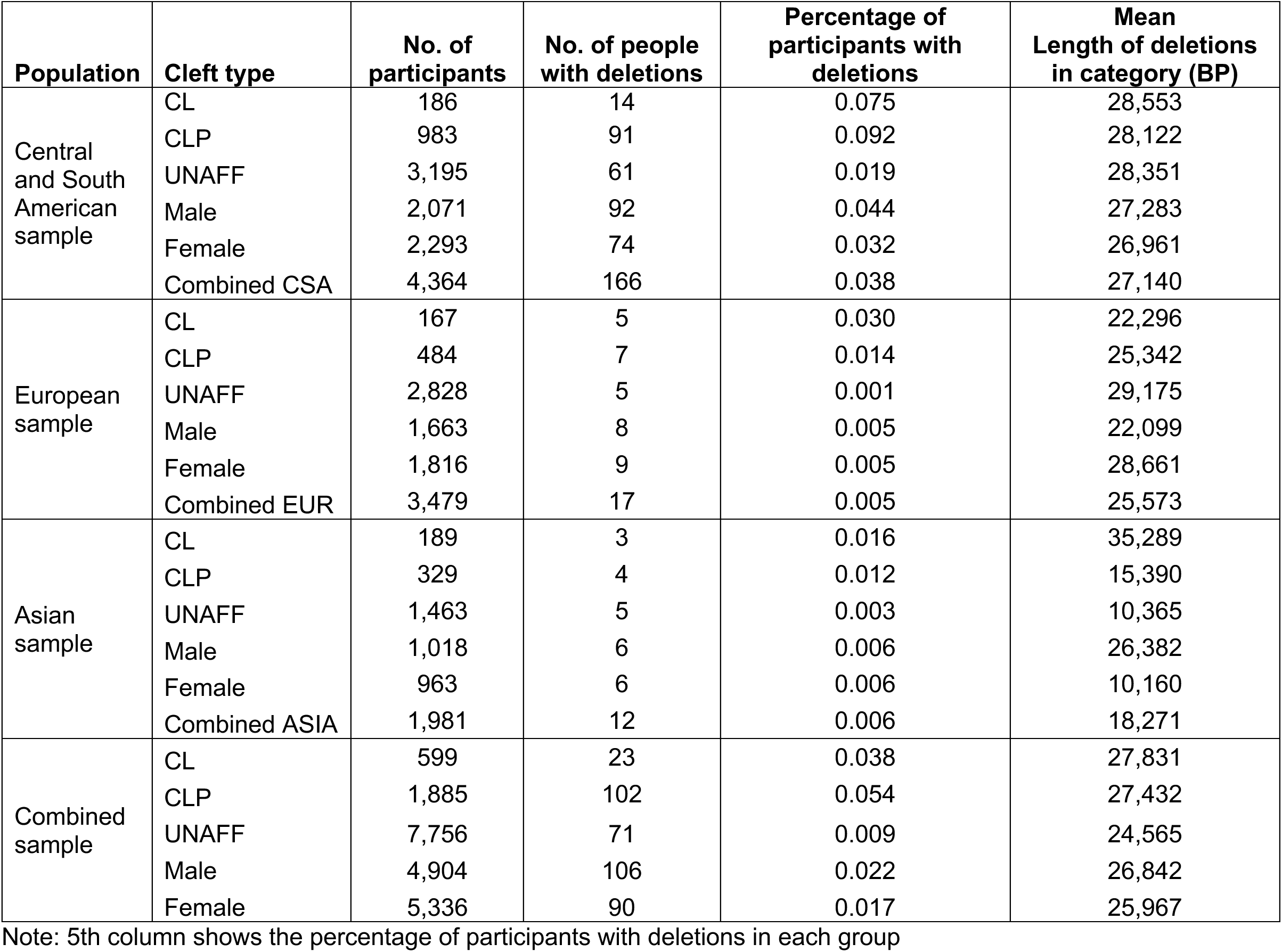
Distribution of deletions by affection status, ancestry and cleft type in 7p14.1 locus.

| **Population** | **Cleft type** | **No. of participants** | **No. of people with deletions** | **Percentage of participants with deletions** | **Mean Length of deletions in category (BP)** |
| --- | --- | --- | --- | --- | --- |
| Central and South American sample | CL | 186 | 14 | 0.075 | 28,553 |
|  | CLP | 983 | 91 | 0.092 | 28,122 |
|  | UNAFF | 3,195 | 61 | 0.019 | 28,351 |
|  | Male | 2,071 | 92 | 0.044 | 27,283 |
|  | Female | 2,293 | 74 | 0.032 | 26,961 |
|  | Combined CSA | 4,364 | 166 | 0.038 | 27,140 |
| European sample | CL | 167 | 5 | 0.030 | 22,296 |
|  | CLP | 484 | 7 | 0.014 | 25,342 |
|  | UNAFF | 2,828 | 5 | 0.001 | 29,175 |
|  | Male | 1,663 | 8 | 0.005 | 22,099 |
|  | Female | 1,816 | 9 | 0.005 | 28,661 |
|  | Combined EUR | 3,479 | 17 | 0.005 | 25,573 |
| Asian sample | CL | 189 | 3 | 0.016 | 35,289 |
|  | CLP | 329 | 4 | 0.012 | 15,390 |
|  | UNAFF | 1,463 | 5 | 0.003 | 10,365 |
|  | Male | 1,018 | 6 | 0.006 | 26,382 |
|  | Female | 963 | 6 | 0.006 | 10,160 |
|  | Combined ASIA | 1,981 | 12 | 0.006 | 18,271 |
| Combined sample | CL | 599 | 23 | 0.038 | 27,831 |
|  | CLP | 1,885 | 102 | 0.054 | 27,432 |
|  | UNAFF | 7,756 | 71 | 0.009 | 24,565 |
|  | Male | 4,904 | 106 | 0.022 | 26,842 |
|  | Female | 5,336 | 90 | 0.017 | 25,967 |

Note: 5th column shows the percentage of participants with deletions in each group

In the ancestry-specific GWASs, the Central and South American group showed the most significant association (p=1.04e-25), which was consistent with the higher frequency of deletions in this group (8.9% in affected individuals). The European subgroup also showed associated at this locus (p=3.37e-08), but with a lower frequency of deletions (0.5% overall, 1.8% in affected individuals). The Asian ancestry group showed only a weak association (p=0.01) in this region and a low frequency of deletions (0.5% overall). Only a single deletion was called in the African sample. The Central and South American and European groups’ association peaks were located in close proximity to each other; the Central and South American group’s peak observed to be 2,935 bp upstream of the European peak (Figure 3(A), Supplement table 1).

**Figure 3.**
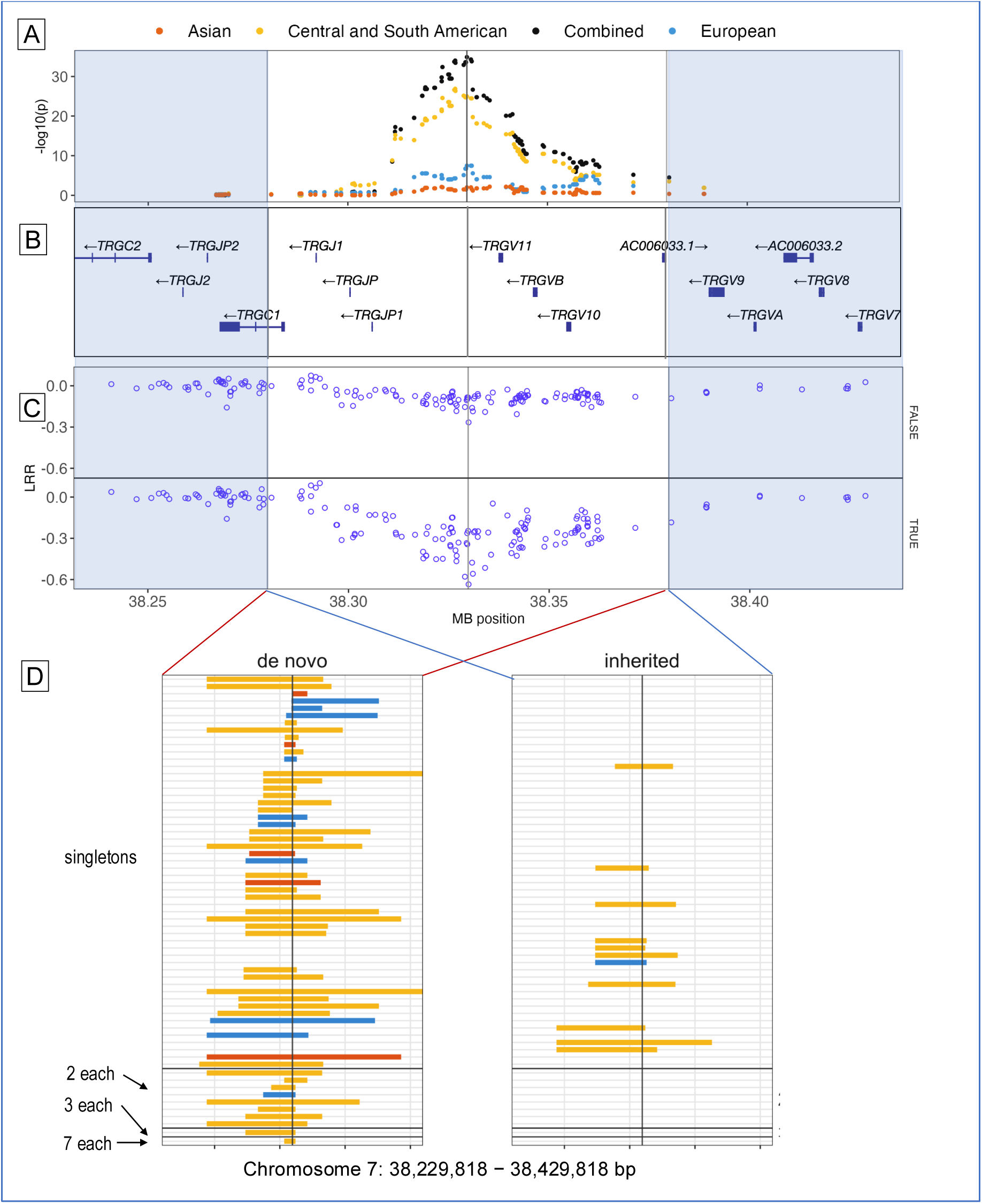
Regional p-value plot of 7p14.1 locus, log R ratios of trios where offspring have de novo deletions, de novo and inherited deletions spanning association peak. Panel (A) regional association (combined and separately by ancestry) and genes located in this region; panel (B) mean LRR of offspring (CL/P and unaffected) with *de novo* deletions, and mean LRR of parents of offspring with de novo deletions at CNV call sites; panel (C) deletions in CL/P individuals spanning the 7p14.1 locus grouped by *de novo* status, ancestry (Central and South American, European and Asian subgroups) and observed frequency.

We also ran a sex-stratified analysis at the 7p14.1 locus, which yielded similar results, indicating no sex difference. Deletions at this locus were on average longer in affected individuals (mean 27,505 bp) than unaffected individuals (mean 24,565 bp), but there were no systematic differences in CNV length by cleft type, ancestry group, or affection status.

The 100 kb region of 7p14.1 spanned by deletions in our study overlaps a 62 kb region of *de novo* deletions reported by Younkin et al (Younkin et al., 2014) as being associated with OFCs, also subsequently replicated in a German case-control sample (Klamt et al., 2016). Both studies reported a higher frequency of deletions in CL/P affected individuals compared to unaffected individuals. The most significant difference between the results from our study and the two previous studies is that we confirmed the association in a combined study sample of participants from multiple ancestries, not only those of European ancestry. More notably, the Central and South American group showed a stronger association as compared to the European group in our study. We also observed that the proportion of affected individuals with DELs in this region was approximately five times as high as the proportion of unaffected individuals (Figure 3(C) and Table 3). The European study (Younkin et al., 2014) observed an approximately two-fold increase of *de novo* deletions in affected as compared to unaffected participants and the German study (Klamt et al., 2016) reported a more modest increase, approximately 15%.

To provide an external reference for CNV frequencies in this region, we examined the gnomAD structural variant database (version 2.0)(Collins et al., 2020), and the Database of Genomic Variation (MacDonald et al., 2014). The gnomAD database reports two intergenic deletions corresponding to a 100 kb region spanning the peak (38,279,818-38,379 bp): a rare 35 kb deletion between 38,295,995 bp – 38,331,222 bp (frequency 0.06%) and a more common deletion 15 kb deletion between 38,315,910 bp – 38,331,224 bp observed at a frequency of 2%. The rarer 35 kb deletion has been observed only in samples of European (0.1 %) and African (0.06%) ancestry. The 15 kb deletion has been seen in multiple ancestry groups, most commonly in samples of African ancestry (2.6%) and Admixed American (2.1%) ancestry. In our study sample, 272 participants (6% of Central and South Americans, 1% of Europeans and Asians, and approximately 0.2% of Africans) had deletions overlapping both gnomAD SVs. It is likely that the CNV calls in this region all correspond to the more common shorter deletion but called with a much higher frequency in the Central and South Americans. The database of genetic variation (DGV) reports 26 deletions and 2 duplications spanning this region.

Since the previously reported association in 7p14.1 reported by Younkin et al (Younkin et al., 2014) was with *de novo* deletions, we asked whether our cohort also showed an increased rate of *de novo* deletions. Of the 196 individuals with 7p14.1 deletions, 103 belong to fully genotyped case-parent trios. Of these, 12 trio offspring (11 affected, 1 unaffected) had a parent with an overlapping CNV, and 91 trio offspring (69 affected, 22 unaffected) were considered to have *de novo* deletions because they either did not have a parent with a CNV in this region or the parent’s CNV did not overlap that of the offspring. We visually confirmed the *de novo* deletions by inspecting the mean Log R ratios (LRR), which were decreased two-to-threefold as compared to the mean LRR of their parents at CNV call sites within the peak region as seen in Figure 3(B). A comparison of the LRRs of the 12 parent-offspring pairs with overlapping CNVs showed that parents’ LRRs were similar to their offspring (Supplement figure 4). Thus, the majority of our CNVs in this region appear to be *de novo*.

### Concordance with CNV calls in 7p14.1 from next-generation sequencing

We attempted to compare the CNV calls overlapping the 7p14.1 peak position to deletions and duplications (SV) identified with the GATK-SV variant calling pipeline(Collins et al., 2020) in 2,959 of the 10,240 participants from our current study sample, who were also whole genome sequenced. The two methods were expected to differ in the exact positions of called CNVs, so we simply checked whether an individual with a PennCNV called CNV also had an SV deletion or duplication overlapping the same 100 kb region. We found that, overall, there was limited concordance between the callsets: of the 64 individuals with CNVs called by PennCNV, only 35 also had an SV called by gnomadSV. Of these 35, many were of different size or precise location than the PennCNV calls. An additional 1,076 SVs were called in this region that did not correspond to PennCNV calls, although most of these were smaller than what we would expect PennCNV to be able to detect. A larger proportion of the PennCNV-only CNV calls were seen in CL/P-affected individuals than in the unaffected individuals, whereas the SV calls were found in similar proportions of affected and unaffected individuals. Finally, only about 10% of the trio offspring were called with a *de novo* SV, compared to nearly 90% of the trios with PennCNV calls.

The lack of concordance between SV and PennCNV calls and the high frequency of *de novo* CNVs led us to more extensive data checking. We compared whether individuals with CNVs in this region have a higher frequency of CNVs overall, or higher variability of LRR values, as compared to those who have normal copy number. The two groups had similar genome-wide CNV frequency, LRR-SDs and mean absolute deviation of LRRs, therefore, the high *de novo* rate does not appear to be the result of elevated false positive rate in CNV calling. We then examined the LRR and BAF values at call sites in the 7p 14.1 peak region within the 91 trios where only the offspring had a PennCNV called deletion. We found that PennCNV preferentially called deletions when a dip in LRR values at consecutive sites were observed, even if BAF values (and genotype calls) indicated a heterozygotic genotype. This could be interpreted as discrediting the PennCNV calls, though such a conclusion assumes that our visual examination is more reliable than the algorithm. We next tested the possibility that the CNV calls are artifacts of plate-related batch effects such as variation in average signal intensities between plates. We flagged plates that showed high deviation in mean LRR-SD, BAF, or total number of called CNVs. However, the *de novo* PennCNV calls do not appear to be artifacts of plate-related variation: only a few of the CNVs (3 out of 31) containing heterozygous genotypes are from outlier plates, and for the majority of these trios with a *de novo* call in the proband (86 out of the 91), the samples of the three family members were plated together.

Thus, these additional investigations did not provide an obvious explanation for the difference between the PennCNV and SV calls. We interpret this at least partially as a demonstration of just how different the two technologies and corresponding algorithms as GATK-SV may not be as tuned for somatic mutations detection as described in the section below.

### An alternative explanation for the 7p14.1 association

Some authors have suggested that T-cell receptor genes such as those in the 7p14.1 show somatic rearrangements within T-cells produced by the thymus throughout the lifespan of a human, the rearrangements being more prevalent in children than adults (van der Velden et al., 2003), and detectable mainly within blood, due to the presence of circulating lymphocytes. It has further been suggested that chip-based genotyping might be more able to detect this effect than sequencing (Sokolowski et al., 2016). We investigated whether this phenomenon could explain our results, particularly in light of the fact that in general our affected individuals are young and our unaffected study subjects are generally older. Because our study includes a mix of both blood and saliva samples, we were able to test for differences between the two. If our results were explained by this hypothesis we would expect to see the “excess” deletions primarily in the youngest individuals, regardless of affection status, and in the blood samples but not saliva. Since saliva has a lower concentration of T-cells than blood, these rearrangements are more likely to be detected in blood samples as compared to the saliva samples.

We examined the deletion frequency within participants whose DNA sample types and ages at time of sample collection were available (9,239 out of the total 10,240). We observed results that are reasonably consistent with the somatic rearrangement hypothesis (figure 4, table 4 and supplement table 2). In blood samples from the Central and South American subgroup, the frequency of deletions is 18% among participants under age 15 with CL/P, 4% among those over age 15 with CL/P, 8% for those under age 15 without CL/P, and under 4% among those over age 15 without CL/P. In saliva samples, all deletion frequencies are 2% or less (table 4).

**Figure 4.**
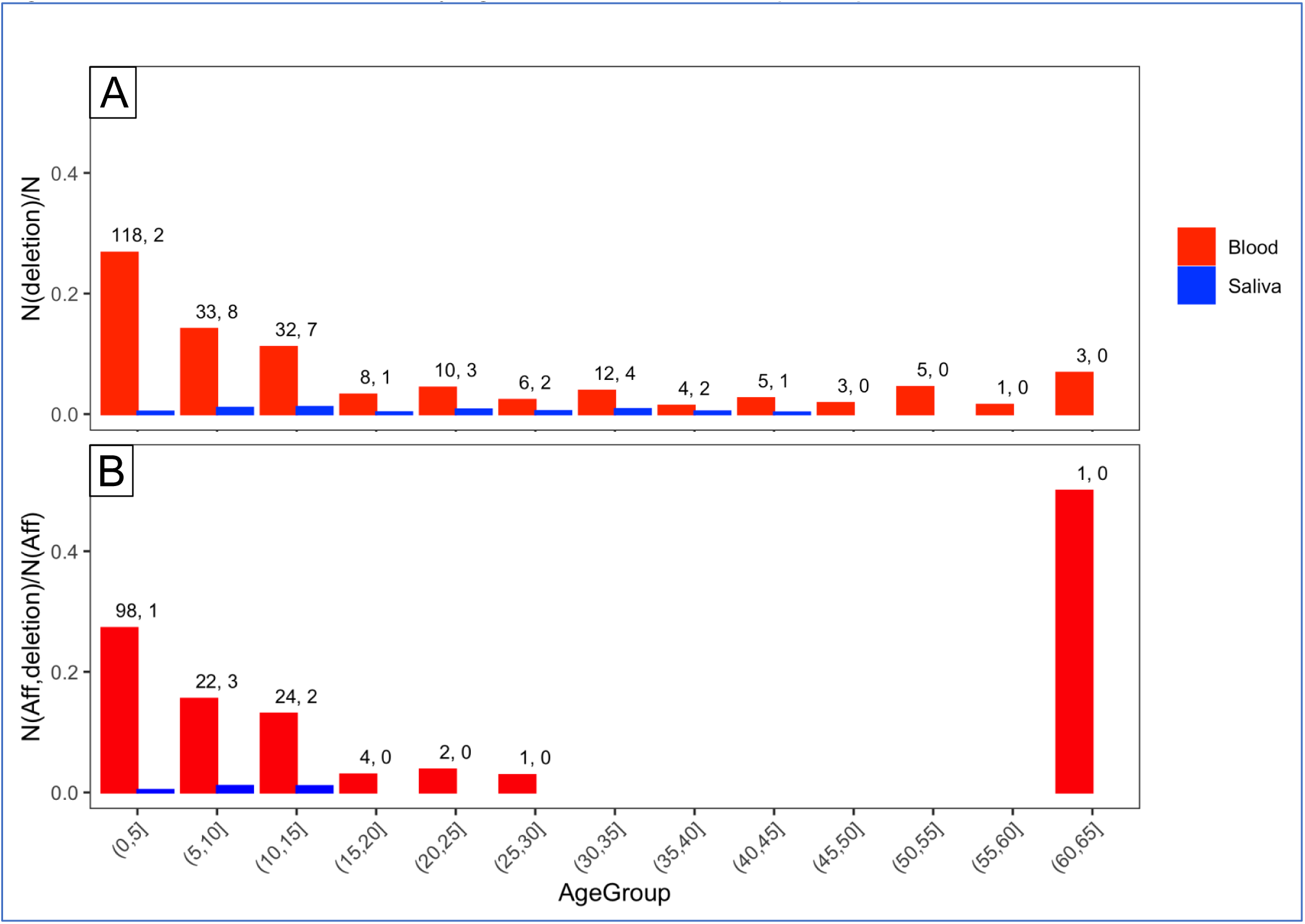
Distribution of deletions by age and DNA source in 7p14.1 peak Deletions seen in complete sample (panel A), and CL/P affected participants (panel B) in a region spanning 38.28 − 38.38 kb on chromosome 7; participants’ ages are grouped into 5 year intervals; numbers above each bar shows number of deletions in blood-derived and saliva-derived samples respectively.

**Table 4.** Frequency of deletions by DNA source and CL/P status in Central and South American ancestry group.

|  | **Blood** |  |  |  | **Saliva** |  |  |  |
| --- | --- | --- | --- | --- | --- | --- | --- | --- |
| **Age** | **CL/P**<br>**Total** | **CL/P with**<br>**deletion**<br>**(%)** | **Unaffected**<br>**Total** | **Unaffected**<br>**with**<br>**deletion**<br>**(%)** | **CL/P**<br>**Total** | **CL/P with**<br>**deletion**<br>**(%)** | **Unaffected**<br>**Total** | **Unaffected**<br>**with**<br>**deletion**<br>**(%)** |
| Under 15 | 735 | 136 (18.5) | 859 | 68 (7.9) | 143 | 1 (0.7) | 334 | 7 (2.1) |
| >=15 | 211 | 8 (3.8) | 1,167 | 41 (3.5) | 42 | 0 | 780 | 5 (0.6) |

Note, however, that a simple Chi-squared test for association between CL/P and presence of deletions remains statistically significant even when restricted to the under-15 group and the blood samples (18.5% vs. 7.9%, p = 0.04, supplement table 2). Thus, there is some ambiguity in interpretation of these results – they could reasonably be interpreted to support either the hypothesis of a true genetic association or the somatic rearrangement hypothesis.

## DISCUSSION

We conducted a genome-wide study to assess whether abnormal copy number is associated with the risk of developing CL/P. Our approach was to test for association of copy number differences at discrete positions in the genome, rather than genotyping individual deletions or duplications, which can require more precise delineation of breakpoints and distinguishing homozygous from heterozygous copy number changes. This approach is fast and flexible, allowing a way to test deletions and duplications together or separately and largely irrespective of CNV length.

Previous studies of copy number variants in OFCs have employed a variety of approaches to call and analyze CNVs including identifying individual microdeletions based on departures from Mendelian inheritance (Shi et al., 2009), array-based comparative genomic hybridization (Lansdon et al., 2023), *de novo* (Younkin et al., 2014) and inherited (Younkin et al., 2015) deletions from SNP arrays, exomes (Ishorst et al., 2024), and cytogenetic arrays (da Silva et al., 2018). The technologies used in these studies vary in CNV detection resolution and scalability; as a result, there have been relatively few large-scale genome-wide assessments of copy number variants in OFCs. Like early GWASs, CNV studies are often limited in diversity and many genetic ancestry groups are not represented.

Because of the differences in study design, it can be difficult to directly compare our results to previous studies. However, we identified a significant association with deletions at the 7p14.1 locus, corroborating a previous association between de novo deletions detected from SNP arrays using a combination of two methods, *MinimumDistance*, and PennCNV. Importantly, the initial association with 7p14 was subsequently replicated in a case-control design in which CNVs were called from SNP arrays using QuantiSNP and then validated with qPCR (Klamt et al., 2016). The observed rate of *de novo* deletions in our study is comparable to that found by Younkin et al. Our study found 91 *de novo* CNVs in 2550 trios (3.6%), the Younkin et al. study found 10 *de novo* CNVs in 467 OFC trios (2.1%), and the difference between the two studies are insignificant (Fisher’s exact test gives p=0.12). Our results build on these discoveries by replicating the association in an independent European population and extending it to a population with admixed American genetic ancestry.

It is perplexing that we observed such extreme inconsistencies between our dataset and the SV calls based on sequence data in a subset of the same individuals. In our cohort, of the 64 participants with PennCNV deletions that were also sequenced, only 35 had a SV call. In addition, the rate of *de novo* deletions called by PennCNV (91 out of 196; 47%) far exceeds the rate of *de novo* SV calls (36 out of 341; 10%). We also found heterozygous SNP genotypes within the regions called deletions by PennCNV, suggesting that at least some of these deletion calls may be inaccurate. One possible explanation is the presence of somatic rearrangements in young individuals, that are detectable mainly within blood, the likelihood of detection being directly proportional to the concentration of circulating lymphocytes. Within our POFC study, CL/P affected participants are typically younger than unaffected participants at the time of sample collection. This is also likely the case with the two previous studies that found an association with 7p14.1 deletions to cleft status (Lansdon et al., 2023; Younkin et al., 2014).

Previous studies of other phenotypes that previously observed associations within TCR regions, e.g. suicidal behavior and type 1 diabetes concluded that these were due to artifacts specific to TCRs (Sokolowski et al., 2016; Zanda et al., 2014). We conclude that the 7p14.1 association with cleft may also be an artifact due to variability in the observed rate of somatic rearrangement deletions by age and DNA source; this variability is also likely responsible for the high frequency of so-called *de novo* deletions observed in our study and the lack of concordance between our genotyping-based CNV calls and sequence-based SV calls.

Except for the 7p14.1 locus, the other loci we report have not been reported in prior CNV studies of OFCs (Conte et al., 2016,; Younkin et al., 2014, 2015). We queried the UCSC genome browser for genes and gene expression related to craniofacial development within 50 kb on either side of each peak’s mid-point as determined by the most significant association p-value. The 1p35.2, 19p13.11 and 21q22.3 loci overlap craniofacial enhancers and active transcription sites, and the 1p42.13 peak overlaps a craniofacial enhancer region. Active transcription start sites are located within 50 kb of the most strongly associated sites in 7p14.1 and 10p12.1. The 7p14.1 locus spans genes from the T cell receptor gamma family, but do not appear to be otherwise connected to embryonic craniofacial development. None of the loci contain genes reported by SNP studies of CL/P, or any OFC in general. Supplement figures 5 a-e show transcription sites and craniofacial enhancer regions for each peak position.

In conclusion, our genome-wide study of CNVs presents an efficient and intuitive approach towards using copy number variation for the discovery of novel genomic regions that control the risk of OFCs using a standard point-by-point GWAS framework. This study also highlights the importance of quality controls and careful analysis while simultaneously making clear the challenges of reconciling disparate datasets and technologies.

## Data Availability

Whole genome genotyping data used to call deletions and duplications, and non-syndromic cleft statuses (including cleft subtype) is available from dbGaP at accession numbers phs000774.v2.p1 and phs000440.v1.p1. Detailed phenotype data, demographics, and medical history is available on FaceBase.org; accession numbers: FB00001368 (doi 10.25550/56-ES6P) and FB00001369 (doi 10.25550/5A-FJBJ).

https://dbgap.ncbi.nlm.nih.gov/home/

https://www.facebase.org/

## ACKNOWLEDGMENTS

The authors wish to thank the participant families worldwide, without whom this research would not have been possible. This work was supported by grants from the National Institutes of Health including:R01-DE016148, R37-DE008559, R01-DE032319, R01-DE031261, R01-DE028300, R01-DE031855, R01-DE032122, X01-HG007845, U01-DE004425, R00-DE024571 and S21-MD001830. Resources for these analyses were also supported in part by the University of Pittsburgh Center for Research Computing and Data (CRCD), RRID: SCR_022735. Specifically, this work used the HTC cluster, which is supported by NIH award number S10-OD028483.

