## Supplemental tables and figures for "Is 7p14.1 an orofacial cleft risk locus? Genome-wide study of copy number variation in multiple populations provides both a replication of previous studies and an alternative explanation"

Supplement Figure 1. Counts of GC-adjusted CNV calls by position within chromosomes 1-22

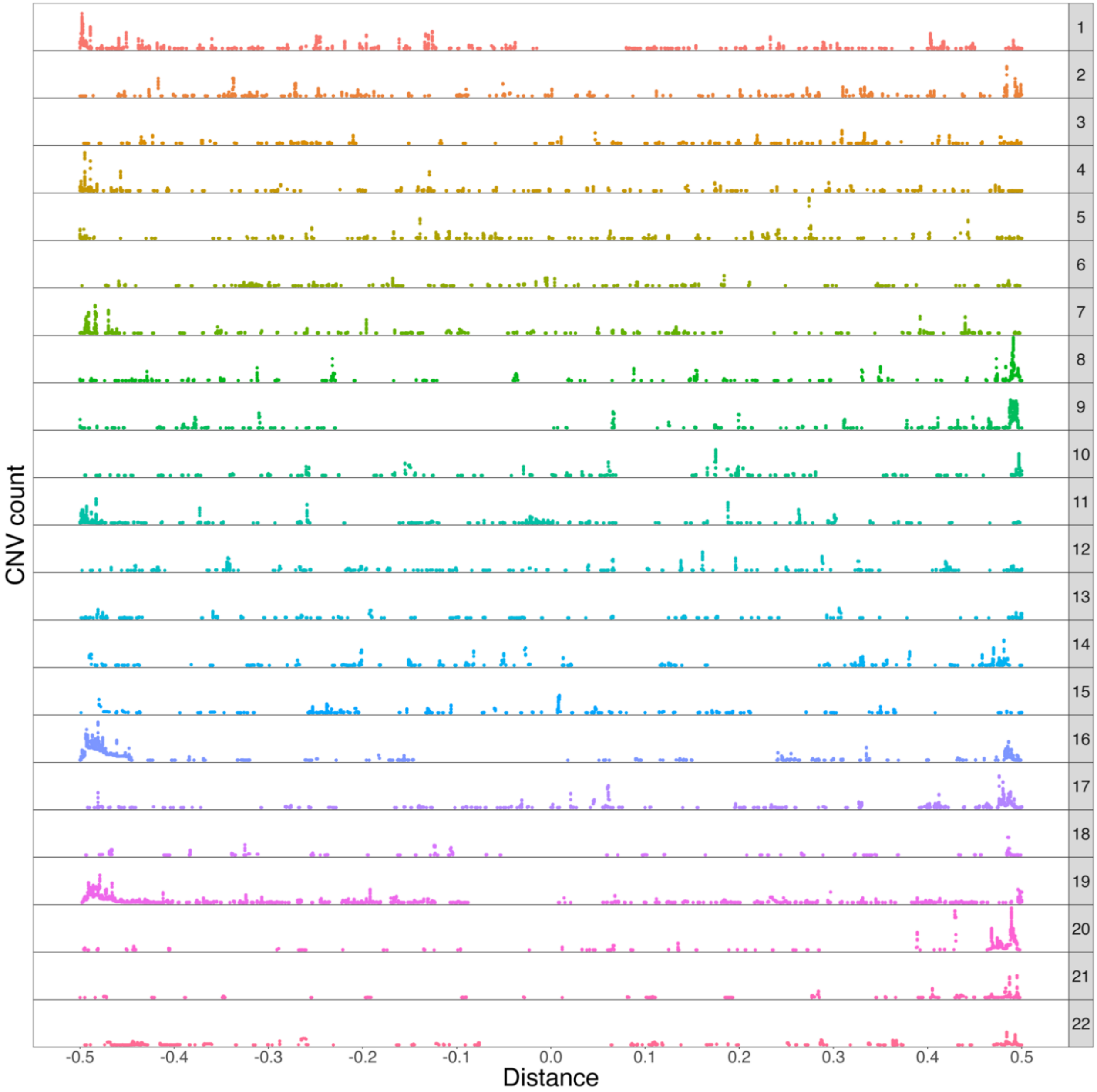


Note. The X-axis shows proportional distance from the midpoint of each chromosome; Y axis values range between 0 and 2000.

Supplement Figure 2: Distribution of CNVs by chromosome

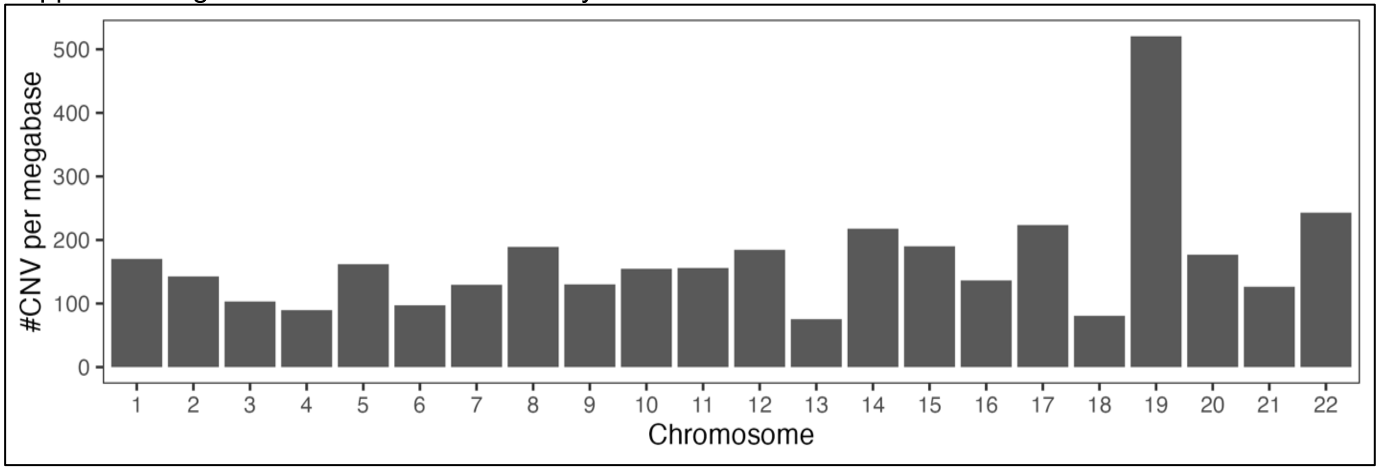

**Supplement table 1. Loci with at least one association p-value < 10e-04 in any ancestry subgroup or TOTAL sample**

| Locus (start MB, end MB) | Asian sample | | European sample | | Central and South American sample | | Combined sample | |
|---|---|---|---|---|---|---|---|---|
| | CNV | DEL | CNV | DEL | CNV | DEL | CNV | DEL |
| 1p35.2 31.7 – 32.7 | 0.062 (32,204,476; 7) | 0.113 (31,905,818; 6) | 0.152 (32,222,088; 8) | **7.55e-05 (32,279,629; 16)[A,B,C]** | 0.071 (31,896,645; 5) | 0.071 (31,896,645; 5) | 0.118 (32,279,629; 342) | 0.167 (32,277,199; 80) |
| 1q42.13 229.0 – 230.0 | 1.48e-04 (229,577,717; 25) | 0.270 (229,623,242; 3) | 0.044 (229,631,673; 26) | 0.076 (229,577,717; 4) | 0.079 (229,619,516; 13) | 0.155 (229,619,516; 6) | **9.72e-05 (229,577,717; 42)[B]** | 0.315 (229,619,516; 19) |
| 7p14.1 37.8-38.8 | 6.61e-03 (38,339,604; 7) | 0.007 (38,339,604; 7) | 3.37e-08 (38,329,818; 17) | 3.37e-08 (38,329,818; 17) | 1.30e-27 (38,326,883; 168) | 1.30e-27 (38,326,883; 168) | **1.32e-35 (38,329,818; 196)[C]** | **1.32e-35 (38,329,818; 196)[C]** |
| 10p12.1 26.8 – 27.8 | 0.036 (27,293,086; 9) | 0.321 (27,382,288; 2) | 8.79e-04 (27,366,407; 46) | 0.004 (27,366,407; 46) | 0.003 (27,524,061; 13) | 0.003 (27,524,061; 13) | **7.07e-05 (27,366,407; 124)[C]** | 0.001 (27,366,407; 71) |
| 12q21.1 74.5 – 75.5 | No CNVs | No CNVs | 0.002 (74,995,693; 15) | 0.002 (74,995,693; 15) | 9.18e-04 (75,054,487; 6) | 0.001 (75,054,487; 6) | **3.86e-05 (74,995,693; 21)** | **3.86e-05 (74,995,693; 21)** |
| 19p13.11 16.5 – 17.5 | 0.125 (16,987,312; 15) | 0.087 (17,008,607; 6) | **1.67e-05 (17,010,308; 12)[B,C]** | 1.30e-04 (17,010,308; 12) | 0.011 (16,987,312; 37) | 0.017 (16,987,312; 37) | 3.42e-04 (17,013,484; 30) | 8.59e-04 (17,013,484; 26) |
| 21q22.3 45.2 – 46.2 | 0.018 (45,825,103; 43) | 0.066 (45815307; 9) | 4.38e-05 (45,773,590; 6) | **1.30e-05 (45,810,795; 6)[A,B,C]** | 0.071 (45,945,534; 7) | 0.025 (45,945,534; 7) | 3.97e-04 (45,825,103; 190) | 0.001 (45,764,397; 25) |
| 22q11.21 20.6 – 21.6 | 0.328 (20,759,825; 9) | 0.179 (20,759,825; 6) | 2.06e-04 (21,119,148; 14) | 1.71e-04 (21,386,010; 14) | 0.004 (21,380,583; 11) | 0.078 (21,342,296; 11) | **4.73e-05 (21,119,148; 24)** | 0.002 (21,386,010; 16) |

Note: For each locus, the lead association is in bold; the smallest p-value seen in other samples within a 100 KB search region spanning the lead association is shown, including p-value, base pair position and number of CNVs and deletions. Base pair position and number of CNVs and deletions are shown in parentheses next to each p-value. For selected associations, **A** indicates the presence of craniofacial super enhancers, **B** indicates craniofacial strong enhancers and **C** indicates transcription site starts locations active during embryonic craniofacial development.

Supplement Figure 3: QQ plots of DEL+DUP and DEL association p-values in Combined sample including 7p14.1 peak and excluding 7p14.1 peak

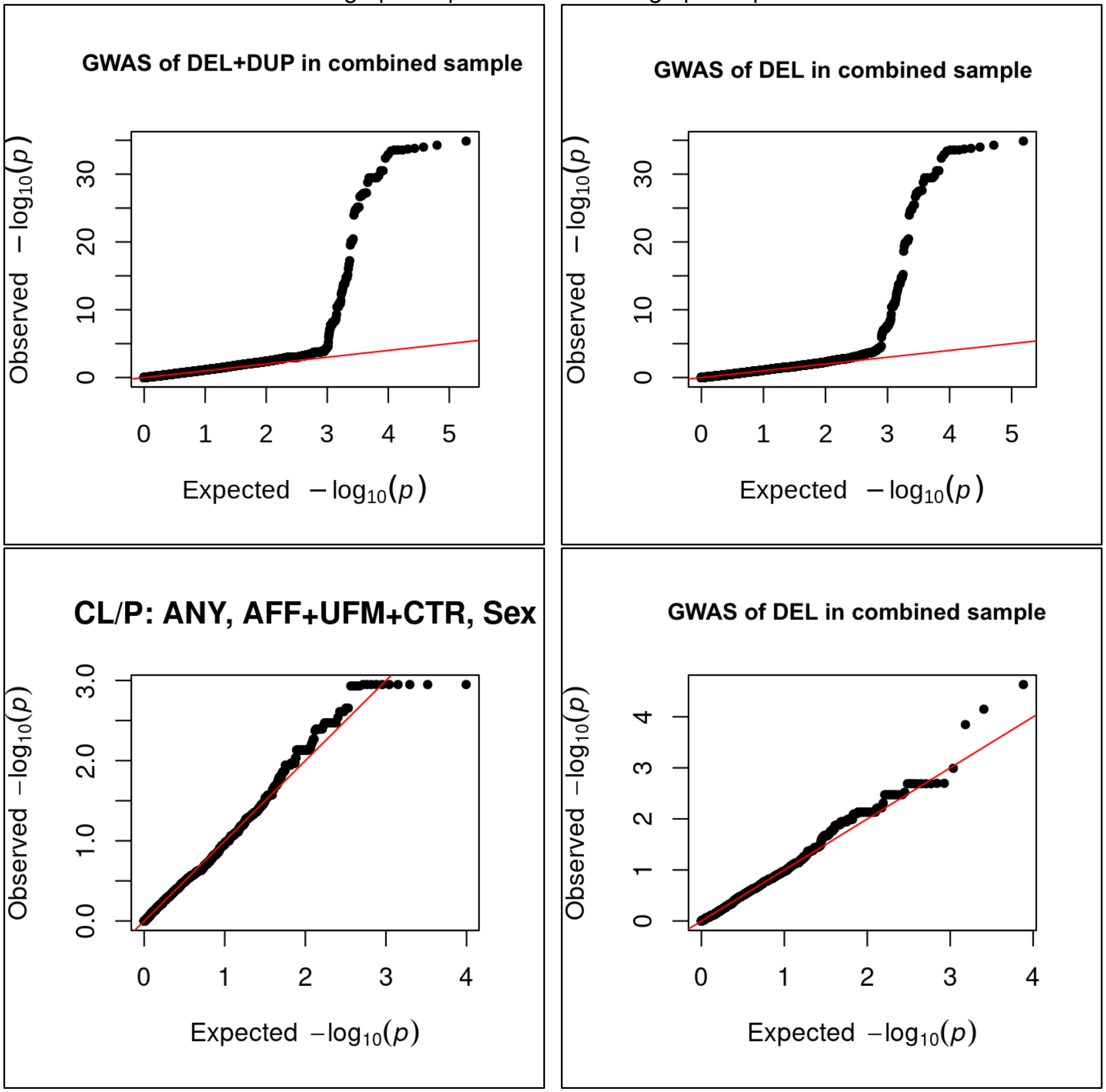

Supplement Figure 4. Parent-offspring pairs with overlapping CNVs in 7p14.1 peak

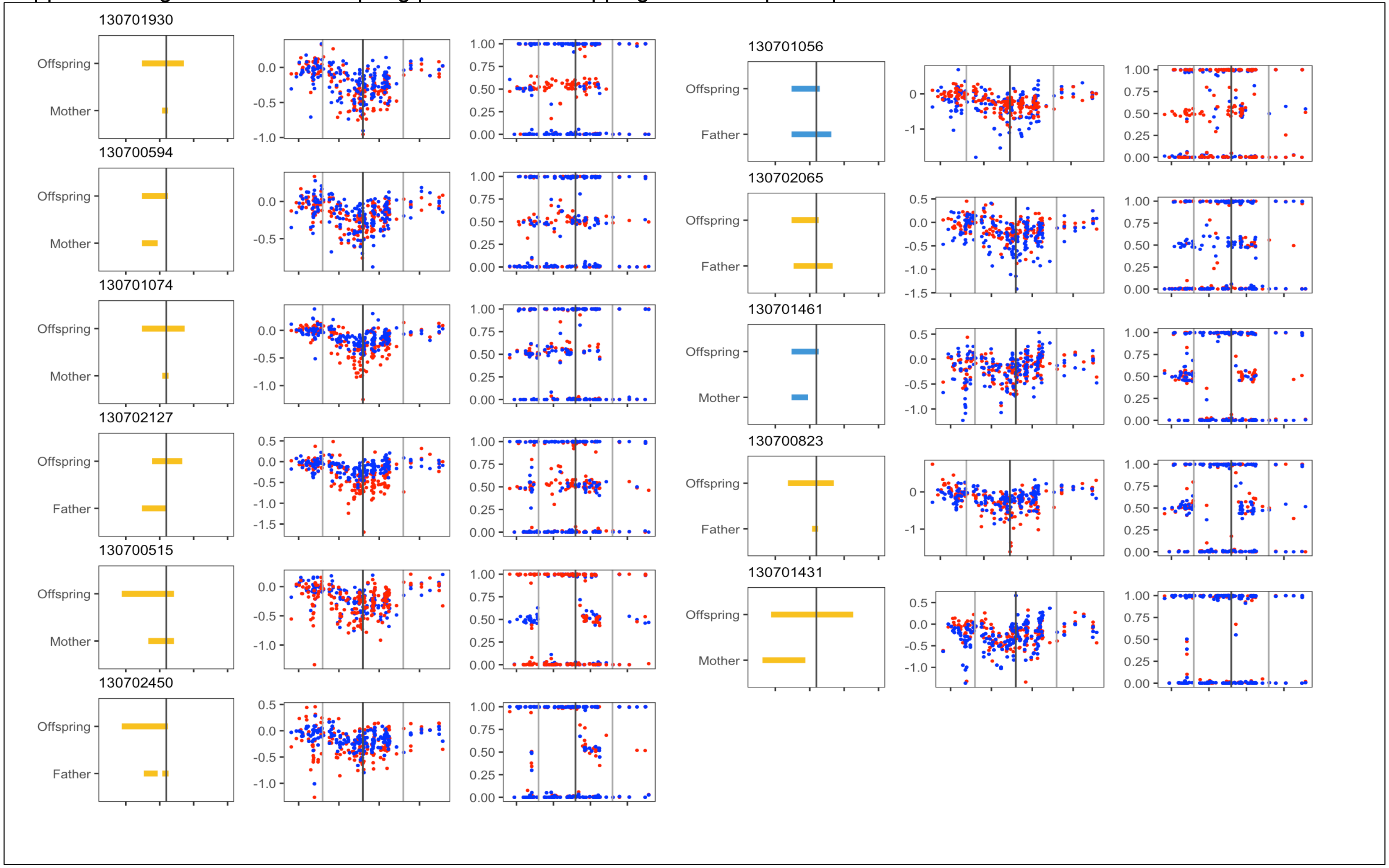


Note: For each pedigree, three plots are shown: (i) deletions, (ii) LRR , and (iii) BAF values at each site used for CNV calling; in the first plot, yellow segments represent individuals of CSA ancestry, and blue, EUR ancestry; in plots 2 and 3, red represents LRR and BAF values of the offspring, and blue the LRR and BAF of parents.

Supplement table 2. Deletions detected in 7p14.1 peak region by DNA source (blood or saliva) and by age

| | | Blood | | | | | | Saliva | | | | | |
|---|---|---|---|---|---|---|---|---|---|---|---|---|---|
| | | CL/P | | Unaffected | | Odds of CL/P in blood sample with deletion | | CL/P | | Unaffected | | Odds of CL/P in saliva sample with deletion | |
| **Ancestry group** | **Age group** | **Total** | **Deleted (%)** | **Total** | **Deleted (%)** | **Odds ratio** | **P-value** | **Total** | **Deleted (%)** | **Total** | **Deleted (%)** | **Odds ratio** | **P-value** |
| **Central and South American** | All | 972 | 145 (14.9) | 2,042 | 111 (5.4) | 3.0 [2.4, 4.0] | 5.8E-17 | 185 | 1 (0.5) | 1,124 | 12 (1.1) | 0.6 [0.0, 3.0] | 1 |
| | Unknown | 26 | 1 (3.8) | 16 | 2 (12.5) | 0.6 [0.2, 1.3] | 0.22 | 0 | 0 | 10 | 0 | NA | NA |
| | Under 15 | 735 | 136 (18.5) | 859 | 68 (7.9) | 1.6 [1.0, 2.4] | 0.04 | 143 | 1 (0.7) | 334 | 7 (2.1) | 0.4 [0.0, 2.2] | 0.44 |
| | >=15 | 211 | 8 (3.8) | 1,167 | 41 (3.5) | 1.0 [0.4, 1.9] | 1 | 42 | 0 | 780 | 5 (0.6) | NA | NA |
| **European** | All | 157 | 15 (9.6) | 503 | 13 (2.6) | 4.0 [1.8, 8.7] | 4.8E-04 | 491 | 3 (0.6) | 2,317 | 11 (0.5) | 1.31 [0.3, 4.3] | 0.73 |
| | Unknown | 42 | 8 (19.0) | 56 | 2 (3.6) | 6.0 [1.4, 45.2] | 0.02 | 25 | 0 | 82 | 0 | NA | NA |
| | Under 15 | 58 | 7 (12.1) | 56 | 6 (10.7) | 1.1 [0.4, 3.9] | 1 | 333 | 3 (0.9) | 663 | 4 (0.6) | 1.5 [0.3, 7.3] | 0.69 |
| | >=15 | 57 | 0 | 391 | 5 (1.3) | NA | NA | 133 | 0 | 1,572 | 7 (0.4) | NA | NA |
| **Combined (incl. Asian and African)** | All | 1,442 | 176 (12.2) | 3,400 | 141 (4.2) | 2.7 [2.1, 3.4] | 2.9E-15 | 1,013 | 6 (0.6) | 4,299 | 24 (0.6) | 1.43 [0.6, 3.1] | 0.37 |
| | Unknown | 369 | 23 (6.2) | 494 | 22 (4.4) | 1.4 [0. 8, 2.6] | 0.28 | 19 | 0 | 33 | 0 | NA | NA |
| | Under 15 | 686 | 144 (20.1) | 277 | 39 (14.1) | 1.6 [1.1, 2.4] | 0.014 | 734 | 6 (0.8) | 1,153 | 11 (1.0) | 0.9 [0.3, 2.3] | 0.81 |
| | >=15 | 387 | 9 (2.3) | 2,629 | 80 (3.0) | 0.7 [0.4, 1.5] | 0.52 | 260 | 0 | 3,113 | 13 (0.4) | NA | NA |

Supplement Figure 5(a) 1p35.2: 32,279,629 ± 50,000 bp

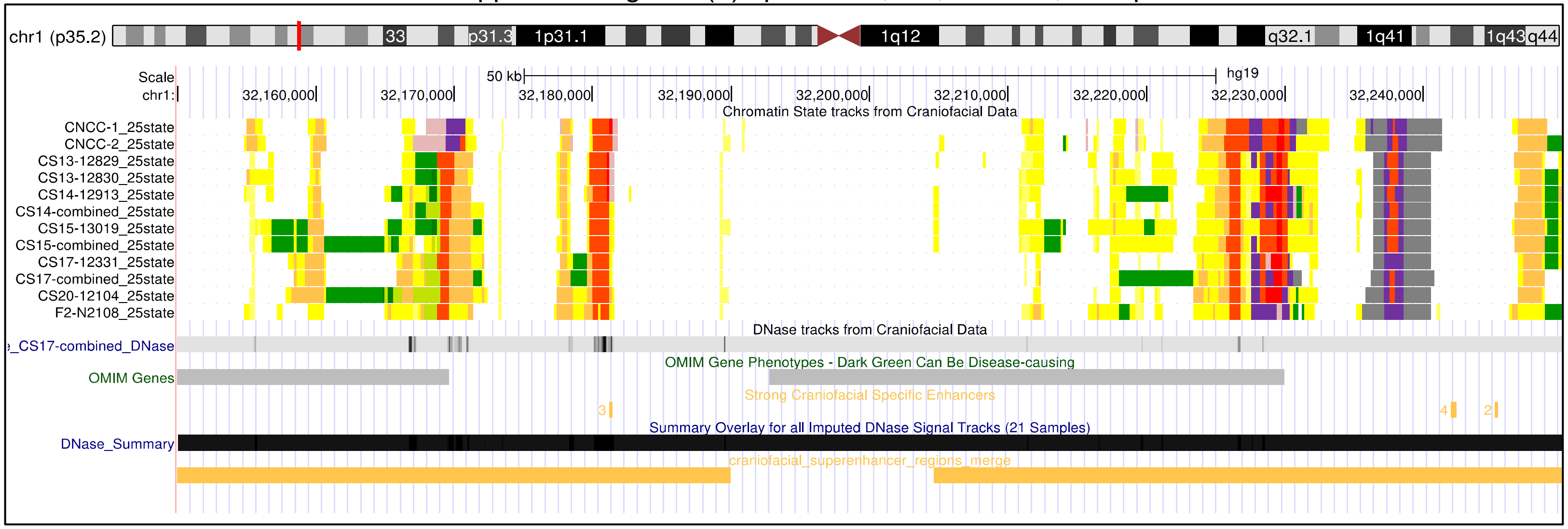


Supplement Figure 5(b) 7p14.1: 38,329,818 ± 50,000 bp

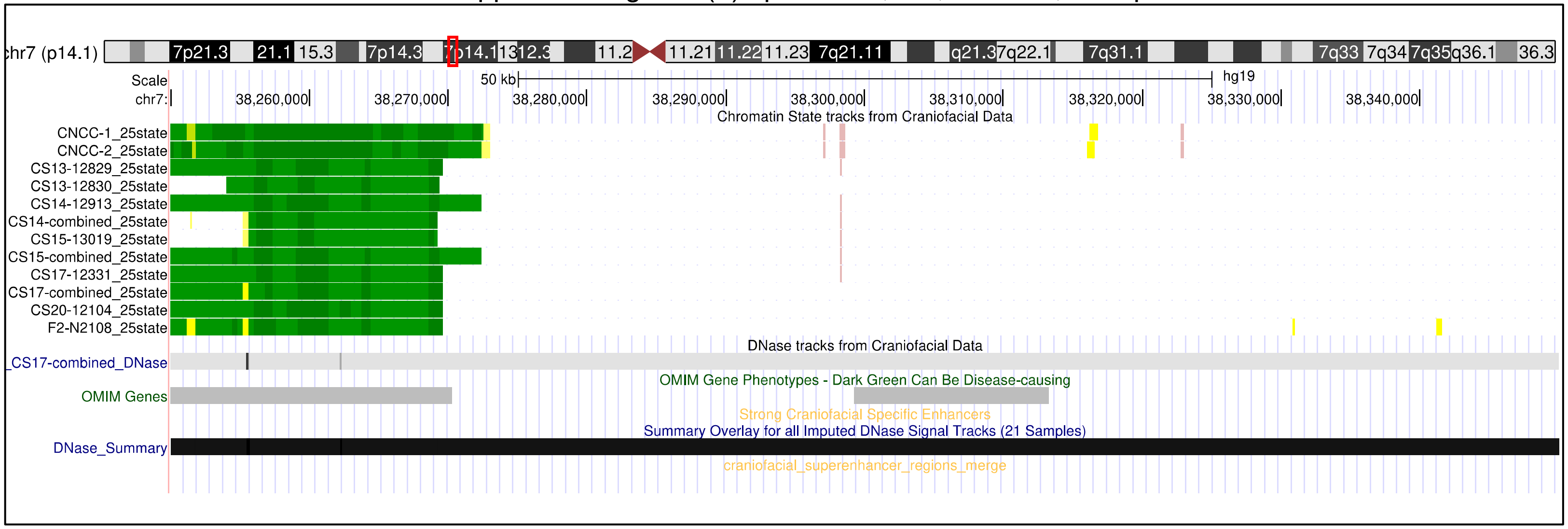

Supplement Figure 5(c) 10p12.1: 38,329,818 ± 50,000 bp

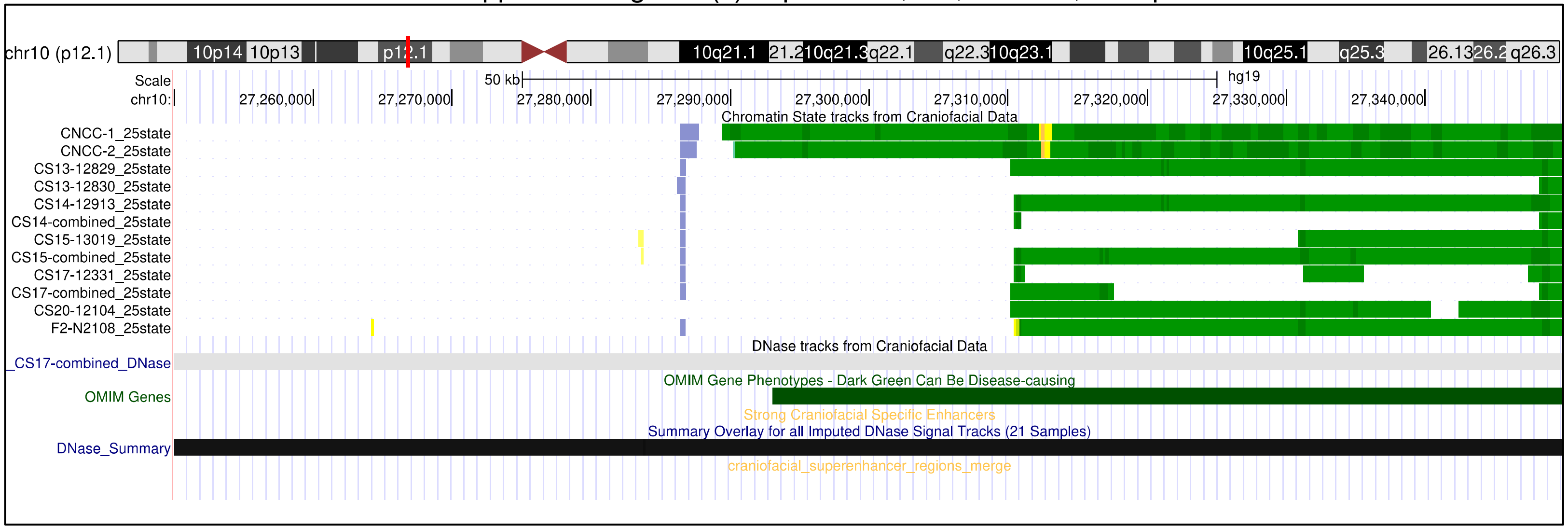


Supplement Figure 5(d) 19p13.11: 17,010,308 ± 50,000 bp

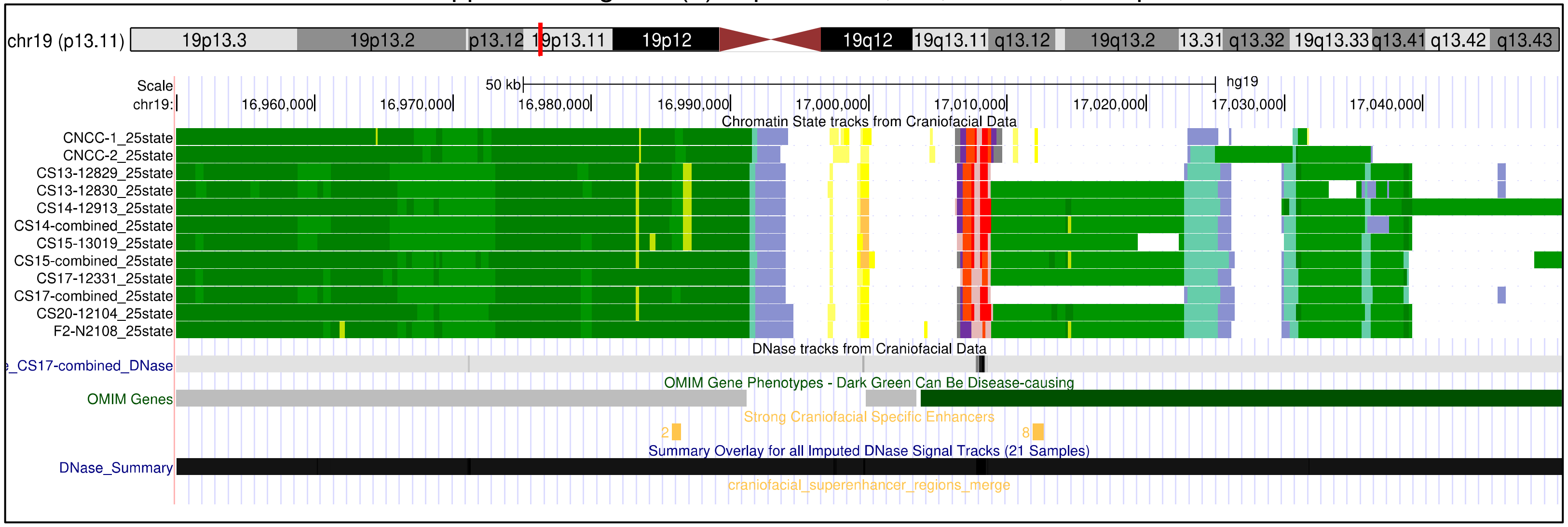

Supplement Figure 5(e) 21q22.3: 45,810,795 ± 50,000 bp

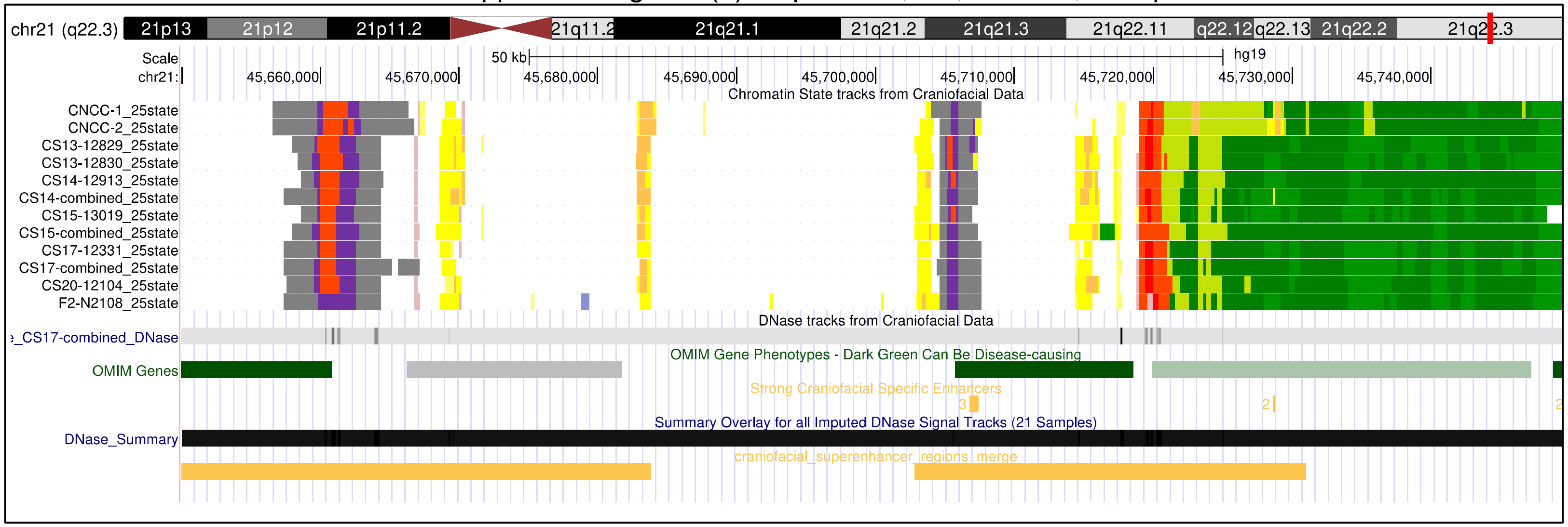
